# Deaths involving COVID-19 by disability status: a retrospective analysis of 29 million adults during the first two waves of the Coronavirus pandemic in England

**DOI:** 10.1101/2021.06.10.21258693

**Authors:** Matthew L. Bosworth, Daniel Ayoubkhani, Vahé Nafilyan, Josephine Foubert, Myer Glickman, Calum Davey, Hannah Kuper

## Abstract

**Objectives:** To assess the association between self-reported disability and deaths involving COVID-19 among adults in England.

**Design:** Cohort study of >29 million adults using data from the Office for National Statistics Public Health Data Asset.

**Setting:** People living in private households or communal establishments (including care homes) in England.

**Participants:** 29,293,845 adults (47% male) aged 30-100 years (mean age = 56) present at the 2011 Census who were alive on 24 January 2020. The main exposure was self-reported disability from the 2011 Census.

**Main outcome measures:** Death involving COVID-19, occurring between 24 January 2020 and 28 February 2021. We estimated the age-standardised mortality rate per 100,000 person-years at-risk, stratified by sex, disability status, and wave of the pandemic. We calculated hazard ratios (HRs) for disabled people compared with non-disabled people, adjusted for geographical factors, socio-demographic characteristics, and pre-pandemic health conditions.

**Results:** Disabled people made up 17% of the study population, including 7% who were ‘more-disabled’ and 10% ‘less-disabled’. From 24 January 2020 to 28 February 2021, 105,213 people died from causes involving COVID-19 in England, 58% of whom were disabled. Age-adjusted analyses showed that, compared to non-disabled people, mortality involving COVID-19 was higher among both more-disabled people (HR=3.05, 95% CI: 2.98 to 3.11 in males; 3.48, 3.41 to 3.56 in females) and less-disabled people (HR=1.88, 95% CI: 1.84 to 1.92 in males; 2.03, 1.98 to 2.08 in females). Among people aged 30-69, HRs reached 8.47 (8.01 to 8.95) among more-disabled females and 5.42 (5.18 to 5.68) for more-disabled males. Sequential adjustment for residence type, geography, socio-demographics, and health conditions partly explained the associations, indicating that a combination of these factors contributed towards the increased risk.

**Conclusion:** Disabled people in England had markedly increased risk of mortality involving COVID-19 compared to non-disabled people and should be prioritised within the pandemic response.

## Background

The COVID-19 pandemic had caused at least 3 million deaths globally by April 2021, including over 127,000 in the UK.^1^ Identifying high-risk groups is critical to target responses, such as vaccine prioritisation. Older people are a key high-risk group, as over 90% of deaths in the UK have been among people aged 60+. ^2^ People with learning disabilities are now recognised as another high-risk group for severe outcomes,^3-11^ in particular people with Down Syndrome.^4 5^ However, evidence is lacking for disabled people more broadly, despite there being at least a billion disabled people globally,^12^ including 11.5 million in England alone.^13^ One exception is a nationwide study in South Korea which found that people with moderate or severe disability were six times more likely to die from COVID-19,^14^ although the total number of deaths was only 228.

There is a strong rationale for an association between disability and COVID-19 mortality. First, any crude association may be due to confounding by age, as disabled people are on average older.^12 13^ Second, disabled people may be clinically vulnerable as they are more likely to have known risk factors for severe COVID-19 (e.g. obesity, diabetes),^15 16^ and the health condition underlying disability may confer increased risk (e.g. Down Syndrome, Parkinson’s Disease).^4 5^ Third, disabled people may be more likely to become infected with COVID-19 as a result of contact with carers, residence in care homes, and/or lack of accessible information on protective measures.^13 17^ Fourth, outcomes may be worse in disabled people if they experience poor quality of treatment or more barriers to accessing care. 18 Fifth, disabled people are more likely to live in poverty and in socio-economically deprived households,^12 13^ 19 which relates to higher risk of COVID-19 death.^16^

People with learning disability are now prioritised for COVID-19 vaccination within the UK, ^20^ as a result of evidence from prospective studies.^5^ Residents in care homes for older adults and those considered ‘clinically extremely vulnerable’ from COVID-19 are also prioritised, 21 as is the case across many high-income settings – for instance most USA states. ^22^ While these focused measures will cover a proportion of the disabled population, other disabled people are not explicitly prioritised for vaccination in the UK. The UK is on track to vaccinate all adults by July 2021, and so considering a policy change on prioritisation may not seem necessary. However, the evidence regarding prioritisation of disabled people is relevant for any future waves of the pandemic (including emerging variants), potentially for policy on other infectious diseases, and for other countries.

In this study, we use population-level data from England, containing detailed socio-demographic characteristics and information on pre-pandemic health status to estimate the association of death involving COVID-19 with self-reported disability, building upon a previous report. 23 We explore whether risk varies between the first and second “waves” of the pandemic, and the likely reasons for any associations.

## Methods

### Study design and data

We conducted a retrospective cohort study of adults aged 30 to 100 years living in private households or communal establishments (including care homes) in England, using data from the Office for National Statistics (ONS) Public Health Data Asset (PHDA). The PHDA comprises linked data from the 2011 Census, General Practice Extraction Service (GPES) Data for Pandemic Planning and Research (GDPPR), ^24^ Hospital Episode Statistics (HES) Admitted Patient Care (APC), 25 and death registrations. The GDPPR dataset contains primary care records for NHS patients with active current registrations at participating practices at the start of the pandemic. Hence, the study population includes people enumerated at the 2011 Census who were alive on 24 January 2020 and could be linked to the 2011 to 2013 Patient Registers and GDPPR dataset. We excluded individuals aged less than 30 years in 2020 from the study population, as their living circumstances are likely to have changed since 2011.

### Outcome

The outcome of interest was death involving COVID-19 (i.e. COVID-19 ICD-10 code of U07.1 or U07.2 anywhere on the death certificate) during the period 24 January 2020 (the date when the first COVID-19 case was reported in the UK) ^26^ and 28 February 2021.

### Exposure

The exposure of interest was self-reported disability status, retrieved from the 2011 Census question: *“Are your day-to-day activities limited because of a health problem or disability which has lasted, or is expected to last, at least 12 months? Include problems related to old age”*. The response options were “Yes, limited a lot” (classified as ‘more-disabled’), “Yes, limited a little” (‘less-disabled’) and “No” (‘non-disabled’).

### Covariates

The following covariates were included, derived from different sources of data:

- Census data (collected 2011): age, residence type, household tenure, National Statistics Socio-Economic Classification (NS-SEC) of household reference person, level of highest qualification, ethnicity, household size, family type, household composition, key worker (individual), key worker in household.
- GPES (January 2015 to December 2019): body mass index (BMI), chronic kidney disease, cancer and immunosuppression, other health conditions (derived in accordance with the QCOVID risk prediction model). ^26^
- HES APC (April 2017 to December 2019): number of admissions to admitted patient care, number of days spent in admitted patient care.
- Other sources: local authority district (derived from the National Statistics Postcode Lookup), local population density, 27 and index of multiple deprivation (IMD) ^28^ (according to postcodes from GPES); individual and household exposure to disease and proximity to others (according to 2011 Census occupation, see Table 1 footnotes for more information); care home residence status (2019 NHS Patient Register).

### Statistical analyses

Characteristics of the study population (means for continuous variables and proportions for categorical variables) were compared across disability groups using standardised differences (*d*), where *d* > 0.1 indicated a large difference between groups.

**Table 1.** Demographic and clinical characteristics for the study population, stratified by sex and disability status

| Variable | Males |  |  | Females |  |  |
| --- | --- | --- | --- | --- | --- | --- |
|  | Non disabled | Less-disabled | More-disabled | Non disabled | Less-disabled | More-disabled |
| **SOCIO-DEMOGRAPHIC** |  |  |  |  |  |  |
| Age (years) Mean (SD) | 54.1 (14.8) | 66.3 (15.5) | 64.6 (15.0) | 54.1 (15.2) | 67.7 (16.0) | 67.3 (15.7) |
| **Place of residence** |  |  |  |  |  |  |
| Private household | 99.3% | 97.5% | 95.5% | 99.4% | 96.9% | 94.5% |
| Care home | 0.2% | 1.7% | 3.2% | 0.4% | 2.8% | 4.9% |
| Other communal establishments | 0.5% | 0.8% | 1.3% | 0.2% | 0.3% | 0.6% |
| **Ethnicity** |  |  |  |  |  |  |
| Bangladeshi | 0.7% | 0.6% | 0.7% | 0.6% | 0.7% | 0.8% |
| Black African | 1.4% | 0.6% | 0.7% | 1.6% | 0.8% | 0.9% |
| Black Caribbean | 1.0% | 1.0% | 1.2% | 1.1% | 1.2% | 1.4% |
| Chinese | 0.6% | 0.3% | 0.2% | 0.6% | 0.3% | 0.2% |
| Indian | 2.9% | 2.1% | 2.1% | 2.7% | 2.3% | 2.7% |
| Mixed | 1.2% | 0.9% | 1.2% | 1.3% | 1.0% | 1.1% |
| Other | 2.3% | 1.9% | 2.1% | 2.4% | 1.9% | 2.1% |
| Pakistani | 1.8% | 1.7% | 2.0% | 1.6% | 1.8% | 2.3% |
| White British | 83.0% | 87.6% | 86.2% | 82.1% | 86.4% | 84.5% |
|  | Non disabled | Less-disabled | More-disabled | Non disabled | Less-disabled | More-disabled |
| White other | 5.2% | 3.3% | 3.6% | 6.0% | 3.7% | 3.9% |
| **Level of highest qualification** |  |  |  |  |  |  |
| No qualification | 14.2% | 32.5% | 48.4% | 16.5% | 40.0% | 52.5% |
| 1-4 GCSE/O-levels | 13.4% | 11.2% | 10.9% | 14.8% | 12.8% | 12.6% |
| 5+ GCSE/O-levels | 13.4% | 10.5% | 8.6% | 17.1% | 13.7% | 10.9% |
| Apprenticeship | 6.4% | 9.0% | 7.4% | 0.9% | 1.0% | 0.8% |
| 2+ A-levels or equivalent | 13.0% | 9.1% | 6.7% | 12.7% | 7.1% | 5.3% |
| Degree or above | 33.7% | 21.0% | 11.9% | 32.9% | 19.4% | 12.5% |
| Other | 5.8% | 6.7% | 6.2% | 5.0% | 6.0% | 5.3% |
| **Region** |  |  |  |  |  |  |
| East of England | 11.9% | 11.0% | 9.3% | 11.8% | 11.1% | 9.7% |
| East Midlands | 9.0% | 9.6% | 9.3% | 8.9% | 9.4% | 9.1% |
| London | 13.2% | 10.8% | 11.8% | 13.2% | 11.4% | 12.6% |
| North East | 4.7% | 5.9% | 7.4% | 4.8% | 5.8% | 6.7% |
| North West | 12.9% | 14.4% | 17.4% | 12.8% | 14.0% | 16.8% |
| South East | 17.4% | 15.1% | 12.2% | 17.5% | 15.4% | 12.6% |
| South West | 10.7% | 11.1% | 9.5% | 10.8% | 11.0% | 9.5% |
| West Midlands | 10.3% | 11.3% | 11.6% | 10.1% | 11.3% | 11.9% |
|  | Non disabled | Less-disabled | More-disabled | Non disabled | Less-disabled | More-disabled |
| Yorkshire and the Humber | 10.0% | 10.8% | 11.5% | 10.0% | 10.5% | 11.0% |
| Population density (people per square kilometre) Mean (SD) | 3,998.4<br>(4,299.9) | 4,002.1<br>(4,156.8) | 4,461.2<br>(4,409.0) | 3,955.4<br>(4,211.8) | 4,025.9<br>(4,134.0) | 4,465.2<br>(4,391.0) |
| **Index of Multiple Deprivation** |  |  |  |  |  |  |
| Decile 1 (most deprived) | 7.2% | 11.0% | 16.6% | 7.4% | 10.6% | 15.4% |
| Decile 2 | 8.0% | 10.2% | 13.6% | 8.2% | 10.3% | 13.1% |
| Decile 3 | 8.8% | 10.0% | 11.9% | 8.8% | 10.0% | 11.6% |
| Decile 4 | 9.5% | 10.1% | 10.6% | 9.4% | 10.0% | 10.4% |
| Decile 5 | 10.1% | 10.0% | 9.3% | 10.0% | 9.9% | 9.4% |
| Decile 6 | 10.7% | 9.9% | 8.5% | 10.6% | 9.9% | 8.5% |
| Decile 7 | 10.9% | 9.8% | 7.6% | 10.8% | 9.6% | 7.8% |
| Decile 8 | 11.2% | 9.4% | 6.9% | 11.1% | 9.3% | 7.1% |
| Decile 9 | 11.4% | 9.0% | 5.9% | 11.4% | 9.0% | 6.3% |
| Decile 10 (least deprived) | 11.6% | 8.1% | 4.5% | 11.5% | 8.3% | 4.9% |
| Not applicable | 0.7% | 2.5% | 4.5% | 0.6% | 3.1% | 5.5% |
| **HOUSEHOLD CHARACTERISTICS** |  |  |  |  |  |  |
| **Occupation of household reference person** |  |  |  |  |  |  |
| Managerial and professional occupations | 37.4% | 18.2% | 13.5% | 36.5% | 18.1% | 13.4% |
|  | Non disabled | Less-disabled | More-disabled | Non disabled | Less-disabled | More-disabled |
| Intermediate occupations | 21.0% | 15.5% | 14.8% | 21.4% | 14.0% | 13.0% |
| Routine and manual occupations | 28.1% | 26.0% | 34.4% | 27.1% | 24.4% | 29.7% |
| Never worked or long-term unemployed | 1.8% | 4.3% | 6.7% | 2.3% | 4.2% | 6.8% |
| Not applicable | 11.1% | 33.4% | 26.1% | 12.1% | 36.2% | 31.7% |
| Not in a household | 0.7% | 2.5% | 4.5% | 0.6% | 3.1% | 5.5% |
| **Tenure of household** |  |  |  |  |  |  |
| Owned outright | 26.8% | 41.9% | 31.8% | 27.5% | 41.7% | 32.0% |
| Owned with a mortgage | 46.2% | 24.9% | 18.0% | 43.2% | 23.1% | 18.9% |
| Shared ownership | 0.7% | 0.5% | 0.5% | 0.8% | 0.6% | 0.6% |
| Social rented from council | 5.2% | 10.4% | 18.0% | 6.5% | 11.5% | 17.6% |
| Other social rented | 4.5% | 9.0% | 15.3% | 5.7% | 9.7% | 14.6% |
| Private rented | 15.1% | 9.8% | 10.7% | 14.9% | 9.0% | 9.4% |
| Living rent free | 0.8% | 1.0% | 1.2% | 0.8% | 1.2% | 1.5% |
| Not in a household | 0.7% | 2.5% | 4.5% | 0.6% | 3.1% | 5.5% |
| **Household size** |  |  |  |  |  |  |
| 1-2 people | 54.7% | 70.9% | 69.7% | 57.8% | 73.7% | 72.3% |
| 3-4 people | 38.5% | 22.8% | 21.8% | 36.0% | 19.8% | 18.8% |
| 5-6 people | 5.4% | 3.3% | 3.4% | 4.9% | 2.9% | 3.0% |
|  | Non disabled | Less-disabled | More-disabled | Non disabled | Less-disabled | More-disabled |
| 7+ people | 0.7% | 0.5% | 0.5% | 0.6% | 0.5% | 0.5% |
| Not in a household | 0.7% | 2.5% | 4.5% | 0.6% | 3.1% | 5.5% |
| **Family status** |  |  |  |  |  |  |
| Not in a family | 17.0% | 23.2% | 28.2% | 15.4% | 26.1% | 29.1% |
| In a couple family | 77.0% | 68.9% | 60.3% | 70.7% | 58.1% | 50.2% |
| In a lone parent family | 5.3% | 5.4% | 7.0% | 13.3% | 12.7% | 15.2% |
| Not in a household | 0.7% | 2.5% | 4.5% | 0.6% | 3.1% | 5.5% |
| **Household composition** |  |  |  |  |  |  |
| Single-adult household | 14.8% | 26.7% | 31.0% | 17.7% | 34.6% | 36.3% |
| Two-adult household | 39.6% | 44.0% | 38.5% | 37.2% | 37.5% | 34.6% |
| Multi-generational household | 9.4% | 10.4% | 11.0% | 7.5% | 8.1% | 8.5% |
| Other 3+ adults | 13.8% | 7.2% | 7.0% | 11.6% | 6.3% | 6.3% |
| Child in household | 21.6% | 9.1% | 7.9% | 25.4% | 10.5% | 8.9% |
| Not in a household | 0.7% | 2.5% | 4.5% | 0.6% | 3.1% | 5.5% |
| **Keyworker type** |  |  |  |  |  |  |
| Not keyworker | 85.1% | 87.2% | 90.2% | 73.4% | 78.4% | 82.9% |
| Education & childcare | 2.3% | 2.1% | 1.1% | 10.4% | 7.3% | 4.4% |
| Food & necessary goods | 0.8% | 0.9% | 1.0% | 0.5% | 0.7% | 1.0% |
|  | Non disabled | Less-disabled | More-disabled | Non disabled | Less-disabled | More-disabled |
| Health & social care | 3.0% | 2.6% | 2.1% | 11.5% | 10.4% | 9.2% |
| Key public services | 1.4% | 1.1% | 0.8% | 1.9% | 1.3% | 1.0% |
| National & local government | 0.6% | 0.8% | 0.5% | 0.9% | 1.0% | 0.7% |
| Public safety & national security | 2.1% | 1.9% | 1.4% | 0.8% | 0.6% | 0.4% |
| Transport | 2.2% | 2.1% | 2.0% | 0.2% | 0.1% | 0.1% |
| Utilities & communication | 2.5% | 1.4% | 0.9% | 0.5% | 0.2% | 0.1% |
| **Keyworker in household** |  |  |  |  |  |  |
| Yes | 35.1% | 28.2% | 23.1% | 36.8% | 29.2% | 24.5% |
| No | 64.2% | 69.3% | 72.4% | 62.6% | 67.6% | 70.1% |
| Not in a household | 0.7% | 2.5% | 4.5% | 0.6% | 3.1% | 5.5% |
| Individual exposure to disease score (0-100)<br>Mean (SD) | 14.5 (16.7) | 14.5 (15.9) | 13.0 (15.1) | 23.6 (23.6) | 21.8 (22.8) | 19.1 (22.2) |
| Individual proximity to others score (0-100)<br>Mean (SD) | 56.9 (17.5) | 56.3 (19.5) | 52.4 (24.4) | 60.8 (20.6) | 57.1 (24.1) | 51.8 (28.4) |
| Household exposure to disease score (0-100)<br>Mean (SD) | 37.4 (32.4) | 29.4 (27.0) | 27.5 (26.6) | 38.2 (32.2) | 29.2 (25.9) | 27.4 (25.4) |
| Household proximity to others score (0-100)<br>Mean (SD) | 72.3 (18.1) | 67.8 (18.1) | 65.9 (20.9) | 72.2 (18.8) | 66.3 (19.4) | 64.2 (22.2) |
|  | Non disabled | Less-disabled | More-disabled | Non disabled | Less-disabled | More-disabled |
| **HEALTH** |  |  |  |  |  |  |
| **Number of admissions to Admitted Patient Care in past 3 years** |  |  |  |  |  |  |
| 0 | 70.5% | 47.7% | 43.2% | 64.3% | 44.9% | 38.3% |
| 1 | 15.9% | 20.2% | 19.3% | 18.9% | 21.5% | 20.4% |
| 2-3 | 9.3% | 18.6% | 19.7% | 11.9% | 20.3% | 22.1% |
| 4-5 | 2.3% | 7.0% | 8.4% | 2.9% | 7.1% | 9.4% |
| 6-9 | 1.2% | 4.3% | 6.0% | 1.3% | 4.1% | 6.5% |
| 10+ | 0.7% | 2.2% | 3.4% | 0.7% | 2.0% | 3.4% |
| **Number of days spent in Admitted Patient Care in past 3 years** |  |  |  |  |  |  |
| 0 | 91.5% | 80.4% | 76.1% | 88.0% | 78.8% | 73.5% |
| 1 | 4.1% | 8.5% | 10.4% | 5.6% | 8.9% | 11.1% |
| 2-4 | 3.2% | 7.2% | 8.4% | 4.7% | 7.9% | 9.4% |
| 5-9 | 0.8% | 2.3% | 3.0% | 1.1% | 2.7% | 3.5% |
| 10-19 | 0.3% | 0.9% | 1.3% | 0.3% | 1.2% | 1.6% |
| 20-39 | 0.1% | 0.4% | 0.5% | 0.1% | 0.4% | 0.6% |
| 40-69 | <0.1% | 0.1% | 0.2% | <0.1% | 0.1% | 0.2% |
|  | Non disabled | Less-disabled | More-disabled | Non disabled | Less-disabled | More-disabled |
| 70+ | <0.1% | <0.1% | 0.1% | <0.1% | <0.1% | 0.1% |
| **Body mass index (kg/m<sup>2</sup>)** |  |  |  |  |  |  |
| < 18.5 | 0.5% | 0.9% | 1.3% | 1.2% | 1.7% | 2.1% |
| 18.5 to 25 | 16.3% | 15.5% | 14.8% | 22.9% | 17.0% | 14.9% |
| 25 to 30 | 23.6% | 23.9% | 21.6% | 18.8% | 19.4% | 17.6% |
| >= 30 | 15.2% | 21.7% | 25.4% | 17.4% | 25.2% | 30.0% |
| Unknown | 44.4% | 38.0% | 36.9% | 39.6% | 36.6% | 35.3% |
| **Chronic kidney disease (CKD)** |  |  |  |  |  |  |
| No CKD | 98.8% | 95.8% | 95.7% | 98.8% | 95.8% | 95.2% |
| Stage 3 CKD | 1.0% | 3.4% | 3.4% | 1.1% | 3.6% | 3.9% |
| Stage 4 CKD | <0.1% | 0.5% | 0.6% | <0.1% | 0.4% | 0.6% |
| Stage 5 CKD | <0.1% | 0.3% | 0.4% | <0.1% | 0.2% | 0.3% |
| **Cancer & immunosuppression** |  |  |  |  |  |  |
| Blood cancer | 0.9% | 1.9% | 1.9% | 1.0% | 2.0% | 2.0% |
| Respiratory cancer | <0.1% | <0.1% | 0.1% | <0.1% | <0.1% | 0.1% |
| Solid organ transplant | <0.1% | <0.1% | <0.1% | <0.1% | <0.1% | <0.1% |
| Prescribed immunosuppressants | <0.1% | <0.1% | <0.1% | <0.1% | <0.1% | <0.1% |
| Prescribed regular prednisolone | 0.7% | 2.9% | 4.2% | 0.9% | 3.5% | 5.6% |
|  | Non disabled | Less-disabled | More-disabled | Non disabled | Less-disabled | More-disabled |
| Prescribed antileukotriene or long-acting beta2-agonists | 5.1% | 13.7% | 19.4% | 5.9% | 15.9% | 22.7% |
| **Other health conditions** |  |  |  |  |  |  |
| Asthma | 9.6% | 12.9% | 14.6% | 11.6% | 17.9% | 21.7% |
| Atrial fibrillation | 3.4% | 11.1% | 10.3% | 1.9% | 7.9% | 8.6% |
| Chronic obstructive pulmonary disease | 2.4% | 9.9% | 14.4% | 2.0% | 7.8% | 11.9% |
| Cirrhosis of the liver | 0.2% | 0.6% | 1.0% | 0.2% | 0.5% | 0.8% |
| Congestive cardiac failure | 1.4% | 6.5% | 8.0% | 0.7% | 4.1% | 5.8% |
| Coronary heart disease | 5.1% | 18.7% | 20.2% | 1.9% | 9.3% | 12.4% |
| Dementia | 0.7% | 3.2% | 4.0% | 0.9% | 4.6% | 6.1% |
| Diabetes | 8.8% | 24.1% | 27.3% | 6.8% | 18.2% | 23.5% |
| Epilepsy | 0.7% | 2.6% | 5.1% | 0.7% | 2.1% | 4.1% |
| Osteoporotic fracture | <0.1% | 0.1% | 0.1% | <0.1% | 0.4% | 0.5% |
| Parkinson's disease | 0.3% | 1.3% | 1.5% | 0.2% | 0.7% | 1.0% |
| Peripheral vascular disease | 0.8% | 3.9% | 5.2% | 0.4% | 1.9% | 2.7% |
| Pulmonary hypertension or pulmonary fibrosis | 0.3% | 1.2% | 1.6% | 0.2% | 1.1% | 1.7% |
| Rare pulmonary diseases | 0.8% | 2.8% | 3.6% | 0.9% | 2.8% | 3.8% |
| Rare neurological conditions | <0.1% | 0.2% | 0.3% | <0.1% | 0.1% | 0.3% |
|  | Non disabled | Less-disabled | More-disabled | Non disabled | Less-disabled | More-disabled |
| Mental illness | 12.5% | 22.5% | 33.1% | 21.6% | 30.3% | 38.8% |
| Stroke or transient ischaemic attack | 2.3% | 8.1% | 10.3% | 1.7% | 6.7% | 9.3% |
| Venous thromboembolism | <0.1% | <0.1% | 0.1% | <0.1% | <0.1% | <0.1% |
| Rheumatoid arthritis or systemic lupus erythematosus | 0.4% | 1.8% | 2.4% | 0.8% | 3.5% | 5.4% |

Percentages don't add to 100% due to rounding. Keyworker type is defined based on the occupation and industry code. 'Exposure to disease' and 'proximity to others' are derived from the O\*NET database, which collects a range of information about individuals' working conditions and day-to-day tasks of their job. To calculate the proximity and exposure measures, the questions asked were: (i) How physically close to other people are you when you perform your current job? (ii) How often does your current job require that you be exposed to diseases or infection? Scores ranging from 0 (no exposure) to 100 (maximum exposure) were calculated based on these questions using methods previously described by the Office for National Statistics.<sup>45</sup>

We calculated age-standardised mortality rates (ASMRs) for disabled and non-disabled people as deaths per 100,000 person-years at-risk to examine the absolute risk of death involving COVID-19. The age distribution within each group was standardised to the 2013 European Standardised Population. ^29^

We estimated the cumulative incidence of death involving COVID-19 during the analysis period using the Aalen-Johansen estimator to account for the competing risk of death not involving COVID-19. Analyses were adjusted for confounding by age using inverse probability weighting with stabilised weights.

We used Cox proportional hazards models to assess whether differences in the risk of mortality involving COVID-19 by disability status could be accounted for by covariates. We included all individuals who died during the analysis period and a random sample of those who did not, with sampling rates of 5% for disabled people and 1% for non-disabled people; case weights equal to the inverse probability of selection were included in the analysis.

We introduced potential explanatory factors sequentially to see how these affected the relationship between disability and mortality involving COVID-19, as follows:

- Model 1: adjustment for single year of age, included as a second-order polynomial.
- Model 2: additional adjustment for place of residence (private household, care home, and other communal establishments).
- Model 3: additional adjustment for geographic (local authority district and population density of the Lower layer Super Output Area), socioeconomic and demographic factors (ethnicity, highest qualification, IMD decile, household characteristics, key worker status, individual and household exposure to disease, and individual and household proximity to others).
- Model 4: additional adjustment for health status; 1) pre-existing health conditions and BMI 2) number of admissions to and days spent in hospital between April 2017 and December 2019. All the health variables were interacted with a binary indicator that allowed the effects to vary depending on whether the individual was aged 70 years or over.

We explored whether the risk of death involving COVID-19 in disabled people has changed over the course of the pandemic by extending the baseline model to allow for time-dependent age and disability coefficients that varied according to wave of the pandemic. The start of the second wave was defined as 21 August 2020, which corresponds to when the reproduction number for the United Kingdom increased to above 1 (90% confidence interval: 0.9 to 1.1) for the first time since it was first reported (29 May 2020), 30 plus 21 days to allow for a lag between new infections and effects on death rates. Thus, deaths occurring from 12 September 2020 onwards were defined as occurring in the second wave. The follow-up time of people who were still in the study from 12 September 2020 was split into wave one and wave two periods, with wave one outcomes recorded as censored.

All statistical analyses were stratified by sex as the risk of death involving COVID-19 differs markedly by sex.^16^

Missing Census responses were imputed using nearest-neighbour donor imputation, the methodology employed by the Office for National Statistics across all 2011 Census variables. ^31^Disability status was missing in 3.2% of Census returns. BMI data from GPES was converted into a categorical variable and individuals with missing BMI values were placed into an ‘unknown’ category.

All analyses were conducted using R version 3.5.

#### Patient and public involvement

The study was conceptualised, designed, conducted, and reported under the demanding circumstances of the COVID-19 pandemic; hence it was not possible to involve patients or members of the public.

## Results

### Characteristics of the study population

The study included 29,293,845 adults in England, 47% male, aged 30 to 100 years (mean age = 56), 17% of whom reported being disabled on the 2011 Census (7% more-disabled and 10% less-disabled) (Table 1). Mean follow-up time was 397 days (SD = 30.9).

Compared with non-disabled people, disabled people (both groups combined) tended to be older (mean age non-disabled = 54, mean age disabled = 67), were more likely to have no qualifications, have a pre-existing health condition, and have been admitted to hospital in the past three years (Table S1). Disabled people were more likely to live in a care home, or in single-adult households, social rented accommodation, a household where the household reference person was in a non-managerial occupation, and in the most deprived areas.

### Age-standardised rates of deaths involving COVID-19 and cumulative mortality involving COVID-19 by disability status

A total of 527,378 deaths were recorded during the follow-up period, 28% of those being among less-disabled people and 26% among more-disabled people. Of these, there were 105,213 total deaths involving COVID-19 (29% less-disabled; 29% more-disabled), 40,934 deaths involving COVID-19 in wave one (29% less-disabled; 30% more-disabled) and 64,279 deaths involving COVID-19 in wave two (29% less-disabled; 28% more-disabled).

Compared to non-disabled people, the ASMRs for all-cause mortality were substantially higher for both less-disabled and more-disabled groups (Table 2). For death involving COVID-19, the ASMRs for all disabled groups were substantially higher than the non-disabled population for the whole outcome period, and in both waves when considered separately.

**Table 2.** Age-standardised mortality rates per 100,000 person-years at-risk (with 95% confidence intervals) for all deaths and deaths involving COVID-19, stratified by sex, self-reported disability status, and wave of the pandemic.

|  | **All deaths** | **Total COVID-19 deaths** | **Wave 1 COVID-19 deaths<sup>1</sup>** | **Wave 2 COVID-19 deaths<sup>2</sup>** |
| --- | --- | --- | --- | --- |
| **Males** |  |  |  |  |
| Non-disabled | 1413<br>(1405 – 1422) | 291<br>(287 – 295) | 196<br>(192 – 200) | 429<br>(422 – 436) |
| Less-disabled | 2451<br>(2430 – 2472) | 535<br>(526 – 545) | 354<br>(344 – 364) | 800<br>(782 – 818) |
| More-disabled | 3931<br>(3897 – 3965) | 899<br>(883 – 915) | 614<br>(597 – 631) | 1322<br>(1292 – 1352) |
| **Females** |  |  |  |  |
| Non-disabled | 980<br>(974 – 986) | 162<br>(159 – 164) | 102<br>(100 – 105) | 248<br>(243 – 253) |
| Less-disabled | 1681<br>(1666 – 1696) | 318<br>(312 – 324) | 200<br>(194 – 207) | 488<br>(476 – 500) |
| More-disabled | 2973<br>(2946 – 2999) | 627<br>(616 – 639) | 408<br>(395 – 420) | 947<br>(925 – 970) |

<sup>1</sup> 24 January 2020 to 11 September 2020
<sup>2</sup> 12 September 2020 to 28 February 2021

Age-adjusted cumulative mortality involving COVID-19 increased more rapidly for disabled people than non-disabled people at the start of the pandemic and remained consistently higher thereafter (Figure 1). At the end of the outcome period, cumulative mortality involving COVID-19 was 2.99 per 1000 for non-disabled males (95% confidence interval: 2.95 to 3.03), 5.55/1000 for less-disabled males (5.44 to 5.67), and 9.39/1000 for more disabled males (9.20 to 9.59). Among females, these figures were 2.11/1000 (2.08 to 2.15), 3.92/1000 (3.84 to 4.00), and 7.36/1000 (7.20 to 7.52) respectively.

**Figure 1.**
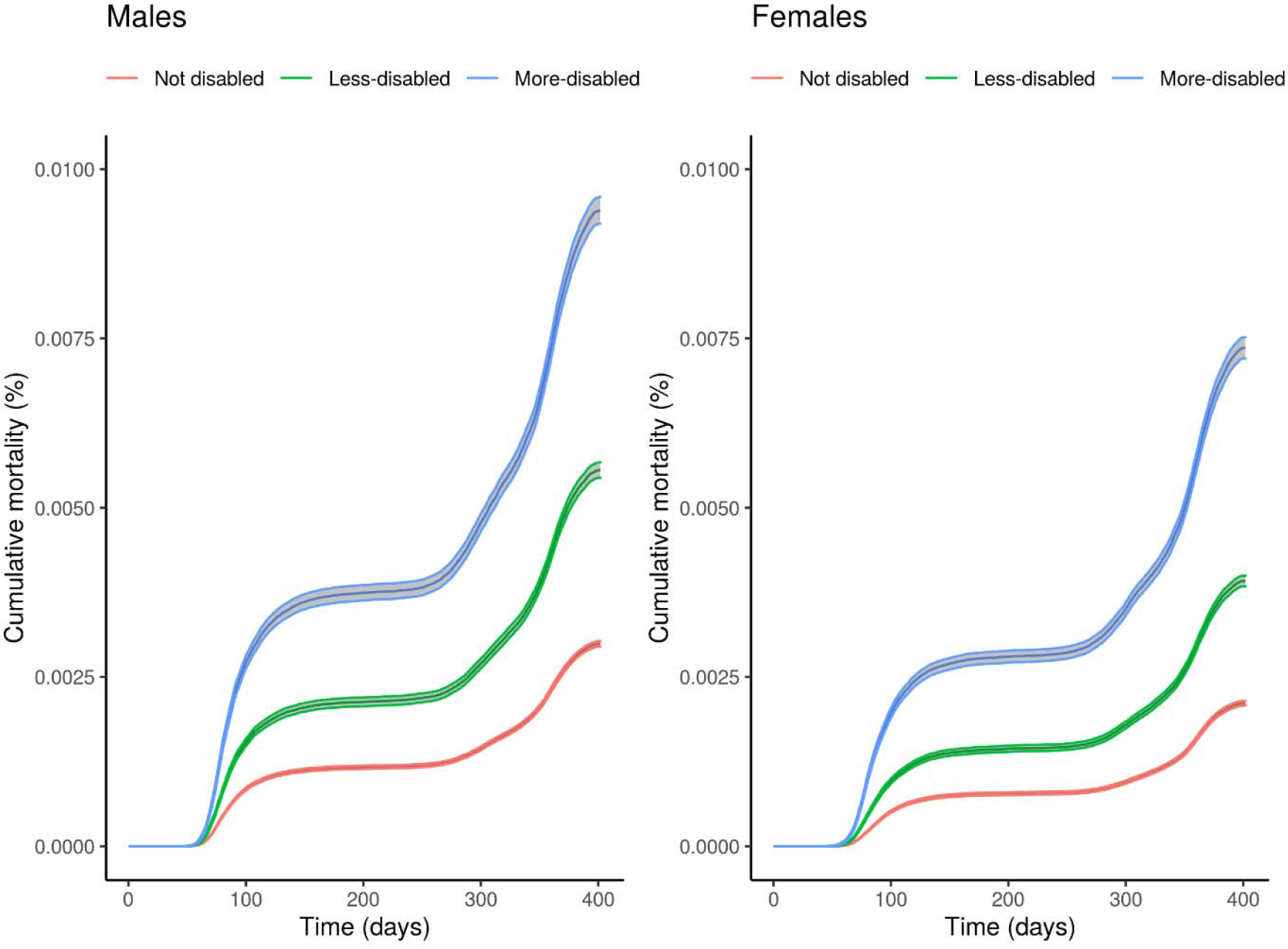
Age-adjusted cumulative COVID-19 mortality during the outcome period (24 January 2020 to 28 February 2021) with bootstrapped 95% confidence intervals by disability status and sex.

### Evaluating the contribution of socioeconomics, demographics, and comorbidities to the excess rates of death involving COVID-19 in disabled people

Consistent with the ASMRs, age-adjusted hazard ratios (HRs) indicated that rates of death involving COVID-19 were substantially higher for both disabled groups compared to non-disabled individuals (Figure 2). Relative to non-disabled males, the rates of death involving COVID-19 were 3.05 (95% CI: 2.98 to 3.11) times greater for more-disabled males and 1.88 (95% CI: 1.84 to 1.92) times greater for less-disabled males. For females, the corresponding HRs were slightly higher at 3.48 (95% CI: 3.41 to 3.56) and 2.03 (95% CI: 1.98 to 2.08), respectively.

**Figure 2.**
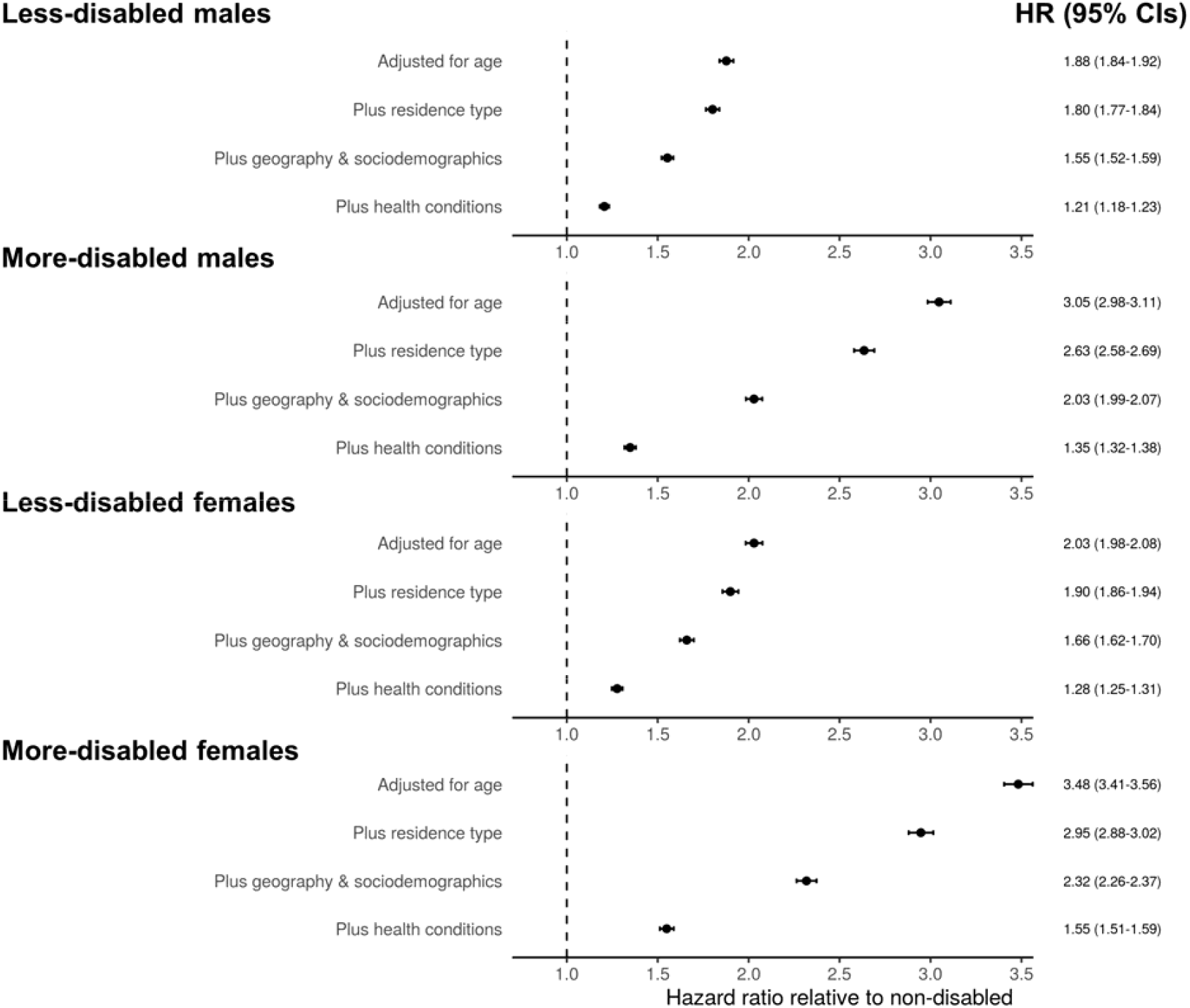
Hazard ratios for deaths involving COVID-19 for less-disabled and more-disabled people relative to non-disabled people, stratified by sex. Results obtained from Cox proportional hazards regression models adjusted for; i) age, ii) plus residence type, iii) plus local authority district, population density, area deprivation, socioeconomic status, ethnicity, household composition, and occupational exposure, and iv) pre-existing conditions. Error bars represent 95% confidence intervals of the hazard ratios.

Including residence type (private household, care home or other communal establishments) in the model partly explained the excess rates of mortality involving COVID-19 for more-disabled people but HRs were largely unchanged for less-disabled people. Additional adjustment for local authority district, population density, and socioeconomic and demographic factors further reduced HRs for all disabled groups. The inclusion of pre-existing health conditions in the model also reduced excess rates of mortality involving COVID-19 further. Across all models, the rate of death involving COVID-19 remained elevated for all disabled groups.

### Potential effect modification of the association of disability and death involving COVID-19: Age, gender, and wave of the pandemic

HRs were substantially higher for all disabled groups aged 30 to 69 years compared with disabled people aged 70 to 100 years, after adjusting for age (Table 3). HRs were higher for women than men for all disabled groups, and this difference was more pronounced in the younger age group. Among people aged 30-69, age-adjusted HRs reached 8.47 (95% CI: 8.01 to 8.95) among more-disabled females and 5.42 (95% CI: 5.18 to 5.68) for more-disabled males compared to their non-disabled counterparts. In the fully-adjusted models, HRs for people aged 30-69 were reduced to 1.91 (95% CI: 1.78 to 2.04) for more-disabled females and 1.74 (95% CI 1.64 to 1.84) for more disabled males (Figure S2).

**Table 3.** Age-standardised mortality rates per 100,000 person-years at-risk (with 95% confidence intervals) and age-adjusted hazard ratios for death involving COVID-19, stratified by age group, sex, and self-reported disability status.

|  | **ASMRs of death**<br>**involving COVID-19** | **Age-adjusted HRs of death**<br>**involving COVID-19** |
| --- | --- | --- |
| **30 to 69 years old in 2020** |  |  |
| *Males* |  |  |
| Non-disabled | 58 (57 – 60) | Reference |
| Less-disabled | 160 (151 – 168) | 2.64 (2.50 – 2.79) |
| More-disabled | 329 (315 – 343) | 5.42 (5.18 – 5.68) |
| *Females* |  |  |
| Non-disabled | 27 (26 – 28) | Reference |
| Less-disabled | 94 (88 – 100) | 3.35 (3.13 – 3.58) |
| More-disabled | 244 (233 – 256) | 8.47 (8.01 – 8.95) |
| **70 to 100 years old in 2020** |  |  |
| *Males* |  |  |
| Non-disabled | 1164 (1147 – 1182) | Reference |
| Less-disabled | 1944 (1911 – 1977) | 1.73 (1.69 – 1.77) |
| More-disabled | 3037 (2982 – 3092) | 2.68 (2.62 – 2.74) |
| *Females* |  |  |
| Non-disabled | 667 (655 – 678) | Reference |
| Less-disabled | 1157 (1137 – 1178) | 1.82 (1.78 – 1.86) |
| More-disabled | 2064 (2026 – 2101) | 2.98 (2.91 – 3.05) |

The increased risk of death involving COVID-19 between disabled and non-disabled people was similar in the first and second waves of the pandemic (Figure 3).

**Figure 3.**
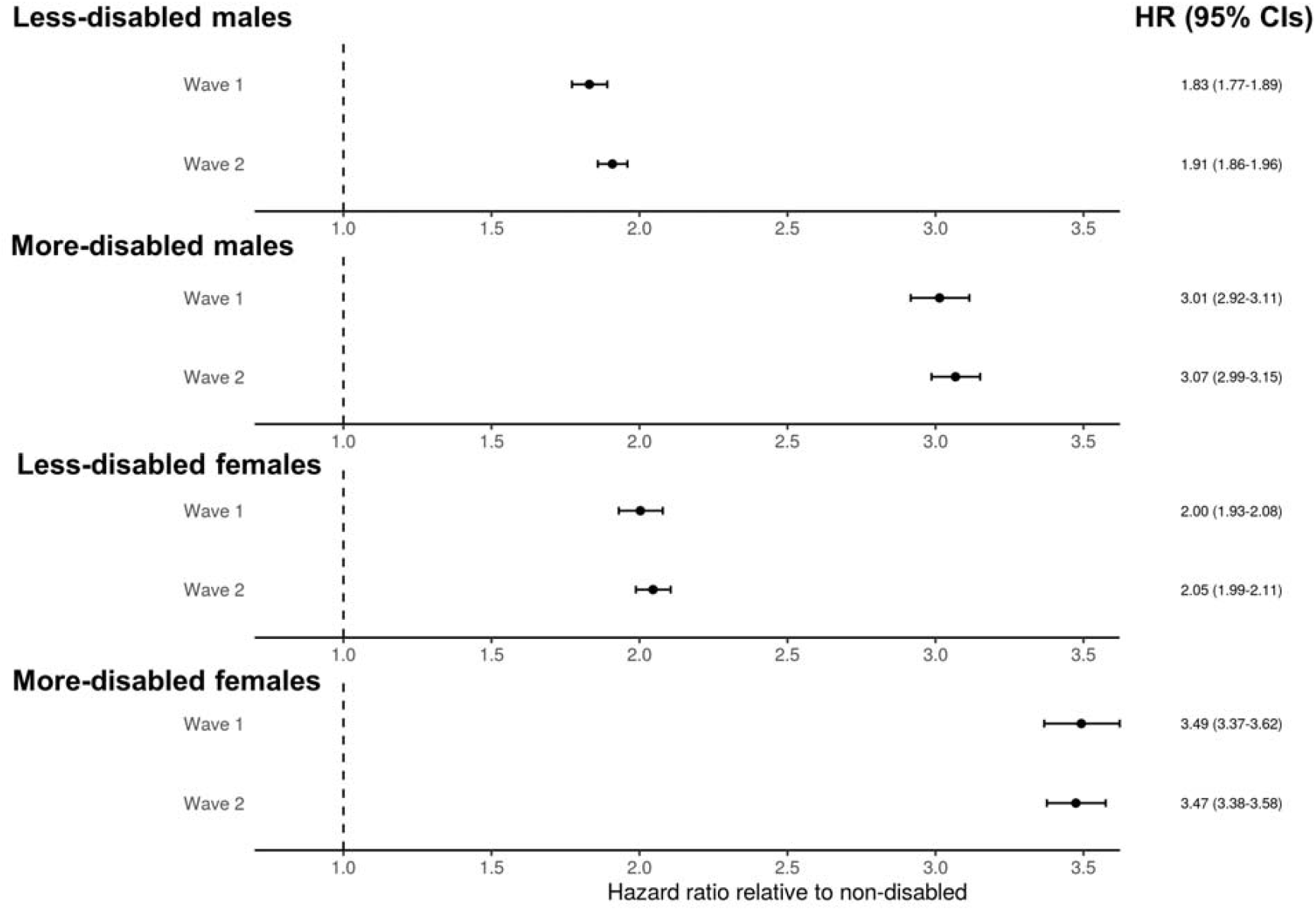
Hazard ratios from the age-adjusted Cox proportional hazards regression model for deaths involving COVID-19 for less-disabled and more-disabled people relative to non-disabled people in wave one (24 January 2020 to 11 September 2020) and wave two (12 September 2020 to 28 February 2021) of the pandemic, stratified by sex. Error bars represent 95% confidence intervals for the hazard ratios.

## Discussion

This large cohort study of 29 million adults in England found that disabled people were at increased risk of death involving COVID-19 compared to non-disabled people. This association was partly explained by adjustment for residence, socio-demographics, geography, and pre-existing health conditions, indicating that a combination of these factors contributed towards the increased risk. Elevated risk of death involving COVID-19 among disabled people was more marked among women compared to men, and younger people (30-69) compared to older (70-100). Indeed, among younger adults, when adjusting for age only, the rate of death involving COVID-19 was 8.5 times greater among more-disabled females and 5.4 times greater for more-disabled males compared to their non-disabled counter-parts. However, the magnitude of the association between disability and COVID-19 mortality was similar for the younger and older age groups in the fully-adjusted models. This suggests that differences in living circumstances and prevalence of pre-existing conditions between younger and older disabled people may mediate this finding. Disabled people were also at excess risk for all causes of death during this period, only approximately 22% of which involved COVID-19 among disabled people in this study population.

## Findings in context

There is clear evidence from the literature that there is an excess risk of COVID-19 mortality associated with learning disability. Analyses using the OpenSAFELY platform showed that people with learning disability were approximately 4-5 times more likely to be hospitalized for COVID-19, and 7-8 times more likely to die, compared to others in the population (with slight variation between waves 1 and 2). ^5^ This link has also been shown by other analyses of medical records in England, including research conducted by the Oxford RCGP Research and Surveillance Centre sentinel network ^8^ and QResearch population level primary care database, ^4^ and by Public Health England, ^9^ using different sources of data. Higher mortality rates among people with learning disability has also been demonstrated in Wales, ^10^ Scotland, ^11^ New York State, ^7^ and the USA more broadly. ^3 6^ Risks are particularly high for people with Down Syndrome. ^4 5^

By contrast, the literature on disability more broadly, or specific impairment types, in relation to COVID-19 outcomes is extremely sparse, and so the current analyses make an important contribution. A nationwide Korean cohort study included 10,237 COVID-19 patients tracked through health insurance records of whom 228 died between January and April 2020. ^14^ The univariable HR for COVID-19 among people with moderate/severe disability compared to those with no disability was 6.2 (4.0-9.7), reducing to 1.6 (1.0-2.6) after extensive adjustment. Univariable associations also showed elevated risks for people with mild disability (4.8, 3.3-6.8), which disappeared after adjustment (1.0, 0.7-1.4). While this evidence is valuable, the sample size was relatively small, variables were adjusted for which are arguably on the causal pathway, and no clear definition was given of disability. Further population-based data of the link between disability overall and COVID-19 mortality could not be identified.

The literature is more extensive with respect to specific conditions that are often disabling. Two systematic reviews highlight the approximately doubled risk of COVID-19 mortality in people with dementia. ^32 33^ A large USA study showed that neurological disorders more broadly was an important predictor of COVID-19 death. ^3^ This finding was supported by evidence from Spain, ^34^ among people with cerebral palsy in the UK, ^26^ and in Scotland among people with multiple sclerosis. ^35^ Studies also show that mortality rates are higher among people who are frail, ^36^ and among nursing home residents with poorer physical function, as assessed through activities of daily living. ^37^ The excess mortality among care home residents is, of course, well documented, and this is particularly high among people with pre-existing conditions such as dementia. ^17 38 39^ International evidence suggests that approximately 41% of COVID-19 deaths in 22 high-income countries were amongst care home residents. ^17^

Evidence is lacking on the link between disability and deaths involving COVID-19, excepting the current study. Most available data on risk factors for COVID-19 is generated through tracking from medical records, often through GP databases. Studies will report risk factors for COVID-19 that are recorded in medical records, such as cardiovascular disease, diabetes, dementia, COPD, cancer, liver and kidney disease, HIV, autoimmune disease, and obesity.^16^ In the UK, learning disability registers, linked to GP records, were established as part of efforts to reduce the health inequalities and poor outcomes experienced by this group. ^40^ However, there is a general failure to record disability or other specific impairment types in medical records. Consequently, it has been possible to identify people with learning disability from GP records and link them to COVID-19 outcomes (although these registers are far from complete) ^5^ but not for disability more broadly.

### Strengths and limitations

The key strength is that the study included >29 million adults and had comprehensive linkage to deaths involving COVID-19. We were able to adjust for a wide range of factors that might confound or mediate the effect of disability sequentially to identify the overall association while also considering possible explanatory factors without over-adjusting the models. These analyses extended previous ONS reports for deaths involving COVID-19 among disabled people which were only until November, 2020, and so missed most of the second ‘wave’. ^23^

Our measure of disability was self-reported and did not include information from clinical records. While this definition relies on an individual’s perception, the measure reflects compliance with the definition of disability within the Equalities Act 2010, which provides a legal basis for protection of disabled people against discrimination and unfair treatment in Great Britain. Hence, the analysis identifies disabled people in a similar way to how disability is identified across policymakers in the UK.

An important limitation is that the measure of disability was from 2011. Consequently, information bias is highly likely, particularly as many older people will have developed disability in the last decade and will consequently be recorded incorrectly as not disabled. In addition, our study population comprised people who were alive at the start of the pandemic but 30% of people who were disabled in 2011 had died before 2020; therefore, our results may be affected by survivorship bias. The most likely result is an under-estimation of the association between disability and mortality, particularly among older people. Moreover, many measures of socio-economic status and demographics were also from the 2011 Census, which will therefore not accurately reflect the situation in 2020. Where possible, this was addressed by using more up-to-date information such as for care home residence. Consequently, there may have been incomplete adjustment, possibly contributing towards the residual association. Information was not collected on disability type, and so it was not possible to disaggregate data to explore which groups were most vulnerable to adverse outcomes. Data was also lacking on key potential mediators, such as healthcare quality and access.

### Policy implications and future research

These analyses demonstrate that disabled people in England are more likely to die from COVID-19, in particular those in younger age groups. As such, they are a high-risk group who could be prioritised in the public health response, including for vaccination. The additional and specific needs of disabled people should be a focus within other activities, such as appropriate and accessible health messaging, testing, shielding, and protection in care homes.

The sequential analysis approach showed the raised risk is because disabled people are disproportionately exposed to a range of generally disadvantageous circumstances, as no single factor explained the results. In addition, we found that disabled people were at higher risk of death from all causes during this period, only a fraction of which involved COVID-19. This finding implies a need to improve services and access to healthcare for disabled people, and tackling the drivers of disadvantage and excess mortality, both during and after the pandemic.

These findings should be replicated and explored further in future studies and in other settings. Administrative data may be the most useful source of information, since existing cohort studies which measure disability may be inadequately powered to show associations with COVID-19 deaths. A number of options could be considered for identifying disability and linking with COVID-19 mortality. Measures of impairment can be generated within the large and comprehensive UK GP registry and used to estimate the association of different impairments with COVID-19 outcomes. However, doing so is time-consuming; requires multi-disciplinary clinical expertise, and identifies people with specific conditions or impairments but not disability as defined in terms of functioning. Another option is to focus on countries where registry data allows linkage of measures of disability (e.g. Swedish disability pension recipients ^41^) or impairment types (e.g. The Finnish Register of Visual Impairment ^42^) to COVID-19 and other health outcomes. As an example, in Sweden a physician-reported registry of adults with rheumatic disease was linked to COVID-19 related deaths. ^43^ A third option is to use health insurance data (or related social security data) to identify people registered with disabilities. ^14^ This approach has been used in the USA across 547 health care organizations, including 64.9 million patients in total, to show that people with intellectual disabilities are at substantially increased risk of COVID-19 mortality. ^3^ Again, this approach relies on disability or specific impairments being recorded within health insurance records. A key need, therefore, is to reach consensus on how recording of disability should be implemented in medical records, ideally aligned with ICF coding. ^44^

### Conclusion

Data from the ONS Public Health Data Asset shows that disabled people were considerably more likely to die from causes involving COVID-19 than non-disabled people of the same age. The analysis suggests that it is a combination of disadvantageous circumstances which explains the increased risk to disabled people. More information is needed to ascertain whether this finding is robust, which protective measures should be put in place (e.g. guidance on shielding and vaccine prioritization), and how policy can tackle mediating factors associated with disability highlighted in this study.

## Data Availability

Information on data availability and access is available via the Secure Research Service: https://www.ons.gov.uk/aboutus/whatwedo/statistics/requestingstatistics/approvedresearcherscheme

## Acknowledgements

HK and CD receive salary support from the PENDA grant from the Foreign, Commonwealth & Development Office.

## Contributors

Contributions are as follows: conceptualisation, MLB, DA, VN, JF, MG, CD, HK; data curation, MLB, VN, DA; formal analysis, MLB; investigation, MLB, DA, VN, JF; methodology, MLB, DA, VN; project administration, MLB, DA, VN, JF, HK; resources, MLB, DA, VN, JF, MG; software, MLB, DA, VN; supervision, MLB, DA, VN, HK; validation, MLB, DA, VN; visualisation, MLB; writing (original draft), MLB, JF, HK; writing (review & editing), MLB, DA, VN, JF, MG, CD, HK. All authors approved the final manuscript. MLB is guarantor for the study.

## Ethical approval

Ethical approval was obtained from the National Statistician’s Data Ethics Advisory Committee (NSDEC(20)12).

## Transparency declaration

The lead author (MLB) affirms that the manuscript is an honest, accurate, and transparent account of the study being reported; that no important aspects of the study have been omitted; and that any discrepancies from the study as planned (and, if relevant, registered) have been explained.

## Dissemination declaration

The size of the study population precludes direct dissemination to participants.

## References

1. Centre for Systems Science and Engineering. COVID-19 Dashboard Baltimore: Johns Hopkins University; 2021 [Available from: https://www.arcgis.com/apps/opsdashboard/index.html#/bda7594740fd40299423467b48e9ecf6 accessed 10.4.2021 2021.

2. Public Health England. COVID-19 confirmed deaths in England (to 31 January 2021): report. London: Public Health England, 2021.

3. Gleason J, Ross W, Fossi A, et al. The Devastating Impact of Covid-19 on Individuals with Intellectual Disabilities in the United States. NEJM Catalyst 2021:March 5

4. Clift AK, Coupland CAC, Keogh RH, et al. COVID-19 Mortality Risk in Down Syndrome: Results From a Cohort Study Of 8 Million Adults. Ann Intern Med 2020 doi: 10.7326/m20-4986 [published Online First: 2020/10/22]

5. Williamson EJ, McDonald HI, Bhaskaran K, et al. OpenSAFELY: Risks of COVID-19 hospital admission and death for people with learning disabilities - a cohort study. medRxiv 2021:2021.03.08.21253112. doi: 10.1101/2021.03.08.21253112

6. Turk MA, Landes SD, Formica MK, et al. Intellectual and developmental disability and COVID-19 case-fatality trends: TriNetX analysis. Disabil Health J 2020;13(3):100942. doi: 10.1016/j.dhjo.2020.100942 [published Online First: 2020/06/01]

7. Landes SD, Turk MA, Formica MK, et al. COVID-19 outcomes among people with intellectual and developmental disability living in residential group homes in New York State. Disabil Health J 2020;13(4):100969. doi: https://doi.org/10.1016/j.dhjo.2020.100969

8. Joy M, Hobbs FR, Bernal JL, et al. Excess mortality in the first COVID pandemic peak: cross-sectional analyses of the impact of age, sex, ethnicity, household size, and long-term conditions in people of known SARS-CoV-2 status in England. Br J Gen Pract 2020;70(701):e890–e98. doi: 10.3399/bjgp20X713393 [published Online First: 2020/10/21]

9. Public Health England. COVID-19: deaths of people with learning disabilities. London: Public Health England, 2020.

10. Improvement Cymru. COVID-19-related deaths in Wales amongst People with Learning Disabilities from 1st March to 26th May 2020. Wales: Improvement Cymru, 2020.

11. Henderson A, Fleming M, Cooper S-A, et al. COVID-19 infection and outcomes in a population-based cohort of 17,173 adults with intellectual disabilities compared with the general population. medRxiv 2021:2021.02.08.21250525. doi: 10.1101/2021.02.08.21250525

12. WHO. World Report on Disability. Geneva: World Health Organisation 2011.

13. Office for National Statistics. Family Resources Survey: financial year 2019 to 2020. London: ONS, 2021.

14. An C, Lim H, Kim DW, et al. Machine learning prediction for mortality of patients diagnosed with COVID-19: a nationwide Korean cohort study. Sci Rep 2020;10(1):18716. doi: 10.1038/s41598-020-75767-2 [published Online First: 2020/11/01]

15. Kinnear D, Morrison J, Allan L, et al. Prevalence of physical conditions and multimorbidity in a cohort of adults with intellectual disabilities with and without Down syndrome: cross-sectional study. BMJ open 2018;8(2):e018292. doi: 10.1136/bmjopen-2017-018292 [published Online First: 2018/02/13]

16. Williamson EJ, Walker AJ, Bhaskaran K, et al. Factors associated with COVID-19-related death using OpenSAFELY. Nature 2020;584(7821):430–36. doi: 10.1038/s41586-020-2521-4 [published Online First: 2020/07/09]

17. Comas-Herrera A, Zalakaín J, Lemmon E, et al. Updated international report: Mortality associated with COVID-19 in care homes, data up to 26th January 2021. UK: International Long Term Care Policy Network, 2021.

18. Sabatello M, Burke TB, McDonald KE, et al. Disability, Ethics, and Health Care in the COVID-19 Pandemic. Am J Public Health 2020;110(10):1523–27. doi: 10.2105/AJPH.2020.305837

19. Pérez-Hernández B, Rubio-Valverde JR, Nusselder WJ, et al. Socioeconomic inequalities in disability in Europe: contribution of behavioral, work-related and living conditions. European journal of public health 2019;29(4):640–47. doi: 10.1093/eurpub/ckz009 [published Online First: 2019/02/13]

20. Public Health England. JCVI advises inviting people on Learning Disability Register for vaccine London: Public Health England; 2021 [Available from: https://www.gov.uk/government/news/jcvi-advises-inviting-people-on-learning-disability-register-for-vaccine.

21. Department of Health & Social Care. Guidance on shielding and protecting people who are clinically extremely vulnerable from COVID-19. London: Department of Health and Social Care, 2021.

22. Johns Hopkins University. Vaccine Prioritization Dashboard Baltimore: Johns Hopkins; 2021 [Available from: https://disabilityhealth.jhu.edu/vaccine/ accessed 10.4.2021 2021.

23. Office for National Statistics. Updated estimates of coronavirus (COVID-19) related deaths by disability status, England: 24 January to 20 November 2020. UK: Office for National Statistics, 2021.

24. NHS digital. General Practice Extraction Service (GPES) Data for pandemic planning and research: a guide for analysts and users of the data. NHS digital, 2020.

25. Herbert A, Wijlaars L, Zylbersztejn A, et al. Data Resource Profile: Hospital Episode Statistics Admitted Patient Care (HES APC). Int J Epidemiol 2017;46(4):1093–93i. doi: 10.1093/ije/dyx015 [published Online First: 2017/03/25]

26. Clift AK, Coupland CAC, Keogh RH, et al. Living risk prediction algorithm (QCOVID) for risk of hospital admission and mortality from coronavirus 19 in adults: national derivation and validation cohort study. Bmj 2020;371:m3731. doi: 10.1136/bmj.m3731 [published Online First: 2020/10/22]

27. Office for National Statistics. Population estimates for the UK, England and Wales, Scotland and Northern Ireland, provisional: mid-2019. Fareham: Office for National Statistics, 2020.

28. Ministry of Housing, Communities & Local Government. English indices of deprivation, 2019. London: Ministry of Housing, Communities & Local Government, 2019.

29. Eurostat. Revision of the European Standard Population. Report of Eurostat’s task force. Luxembourg: European Union, 2013.

30. Department of Health & Social Care and Scientific Advisory Group for Emergencies. Guidance: The R value and growth rate, 2020.

31. Office for National Statistics Census Item Edit and Imputation Process. London: Office for National Statistics, 2012.

32. July J, Pranata R. Prevalence of dementia and its impact on mortality in patients with coronavirus disease 2019: A systematic review and meta-analysis. Geriatr Gerontol Int 2021;21(2):172–77. doi: 10.1111/ggi.14107 [published Online First: 2020/12/20]

33. Hariyanto TI, Putri C, Arisa J, et al. Dementia and outcomes from coronavirus disease 2019 (COVID-19) pneumonia: A systematic review and meta-analysis. Arch Gerontol Geriatr 2021;93:104299. doi: 10.1016/j.archger.2020.104299 [published Online First: 2020/12/08]

34. García-Azorín D, Martínez-Pías E, Trigo J, et al. Neurological Comorbidity Is a Predictor of Death in Covid-19 Disease: A Cohort Study on 576 Patients. Front Neurol 2020;11:781. doi: 10.3389/fneur.2020.00781 [published Online First: 2020/08/01]

35. Fernandes PM, O’Neill M, Kearns PKA, et al. Impact of the first COVID-19 pandemic wave on the Scottish Multiple Sclerosis Register population. Wellcome Open Res 2020;5:276. doi: 10.12688/wellcomeopenres.16349.1 [published Online First: 2021/02/16]

36. Petermann-Rocha F, Hanlon P, Gray SR, et al. Comparison of two different frailty measurements and risk of hospitalisation or death from COVID-19: findings from UK Biobank. BMC Med 2020;18(1):355. doi: 10.1186/s12916-020-01822-4 [published Online First: 2020/11/11]

37. Panagiotou OA, Kosar CM, White EM, et al. Risk Factors Associated With All-Cause 30-Day Mortality in Nursing Home Residents With COVID-19. JAMA Intern Med 2021 doi: 10.1001/jamainternmed.2020.7968 [published Online First: 2021/01/05]

38. Morciano M, Stokes J, Kontopantelis E, et al. Excess mortality for care home residents during the first 23 weeks of the COVID-19 pandemic in England: a national cohort study. BMC Med 2021;19(1):71. doi: 10.1186/s12916-021-01945-2 [published Online First: 2021/03/06]

39. Dutey-Magni PF, Williams H, Jhass A, et al. COVID-19 infection and attributable mortality in UK care homes: Cohort study using active surveillance and electronic records (March-June 2020). Age Ageing 2021 doi: 10.1093/ageing/afab060 [published Online First: 2021/03/13]

40. Heslop P, Blair PS, Fleming P, et al. The Confidential Inquiry into premature deaths of people with intellectual disabilities in the UK: a population-based study. Lancet 2014;383(9920):889–95. doi: 10.1016/s0140-6736(13)62026-7 [published Online First: 2013/12/18]

41. Helgesson M, Rahman S, Saboonchi F, et al. Disability pension and mortality in individuals with specific somatic and mental disorders: examining differences between refugees and Swedish-born individuals. J Epidemiol Community Health 2021 doi: 10.1136/jech-2019-213436 [published Online First: 2021/01/22]

42. Meyer-Rochow VB, Hakko H, Ojamo M, et al. Suicides in Visually Impaired Persons: A Nation-Wide Register-Linked Study from Finland Based on Thirty Years of Data. PLoS One 2015;10(10):e0141583. doi: 10.1371/journal.pone.0141583 [published Online First: 2015/10/29]

43. Strangfeld A, Schäfer M, Gianfrancesco MA, et al. Factors associated with COVID-19-related death in people with rheumatic diseases: results from the COVID-19 Global Rheumatology Alliance physician-reported registry. Ann Rheum Dis 2021 doi: 10.1136/annrheumdis-2020-219498 [published Online First: 2021/01/29]

44. WHO. International Classification of Functioning, Disability and Health. Geneva, Switzerland: WHO, 2001.

45. Office for National Statistics. Which occupations have the highest potential exposure to the coronavirus (COVID-19)? London: Office for National Statistics, 2020.

