## Supplementary Materials for "Deaths involving COVID-19 by disability status: a retrospective analysis of 29 million adults during the first two waves of the Coronavirus pandemic in England"

**Table S1.** Standardised differences comparing people with and without disability for all the covariates included in the modelling

| Variable | Level | *d* |
|---|---|---|
| Age (years) | Mean | 0.82 |
| Place of residence | Private household | 0.16 |
| | Care home | 0.16 |
| | Other communal establishments | 0.03 |
| Ethnicity | Bangladeshi | 0.01 |
| | Black African | 0.06 |
| | Black Caribbean | 0.01 |
| | Chinese | 0.05 |
| | Indian | 0.02 |
| | Mixed | 0.02 |
| | Other | 0.02 |
| | Pakistani | 0.01 |
| | White British | 0.08 |
| | White other | 0.08 |
| Level of highest qualification | No qualification | 0.48 |
| | 1-4 GCSE/O-levels | 0.05 |
| | 5+ GCSE/O-levels | 0.10 |
| | Apprenticeship | 0.02 |
| | 2+ A-levels or equivalent | 0.16 |
| | Degree or above | 0.33 |
| | Other | 0.02 |
| Keyworker type | Not keyworker | 0.10 |
| | Education & childcare | 0.09 |
| | Food & necessary goods | 0.02 |
| | Health & social care | 0.03 |
| | Key public services | 0.04 |
| | National & local government | <0.01 |
| | Public safety & national security | 0.03 |
| | Transport | 0.02 |
| | Utilities & communication | 0.07 |
| Individual exposure to disease score (0-100) | Mean | 0.07 |
| Individual proximity to others score (0-100) | Mean | 0.19 |
| Number of admissions to Admitted Patient Care in past 3 years | 0 | 0.39 |
| | 1 | 0.06 |
| | 2-3 | 0.21 |
| | 4-5 | 0.18 |
| | 6-9 | 0.16 |
| | 10+ | 0.11 |
| Number of days spent in Admitted Patient Care in past 3 years | 0 | 0.26 |
| | 1 | 0.14 |
| | 2-4 | 0.14 |
| | 5-9 | 0.10 |
| | 10-19 | 0.08 |
| | 20-39 | 0.05 |
| | 40-69 | 0.03 |
| | 70+ | 0.02 |

| Variable | Level | *d* |
|---|---|---|
| Body mass index (kg/m$^2$) | < 18.5 | 0.05 |
| | 18.5 to 25 | 0.09 |
| | 25 to 30 | 0.01 |
| | >= 30 | 0.18 |
| | Unknown | 0.09 |
| Chronic kidney disease (CKD) | No CKD | 0.15 |
| | Stage 3 CKD | 0.13 |
| | Stage 4 CKD | 0.06 |
| | Stage 5 CKD | 0.04 |
| Cancer & immunosuppression | Blood cancer | 0.07 |
| | Respiratory cancer | 0.03 |
| | Solid organ transplant | 0.02 |
| | Prescribed immunosuppressants | 0.01 |
| | Prescribed antileukotriene or long-acting beta2-agonists | 0.29 |
| | Prescribed regular prednisolone | 0.15 |
| Other health conditions | Asthma | 0.14 |
| | Atrial fibrillation | 0.22 |
| | Chronic obstructive pulmonary disease | 0.26 |
| | Cirrhosis of the liver | 0.06 |
| | Congestive cardiac failure | 0.19 |
| | Coronary heart disease | 0.29 |
| | Dementia | 0.17 |
| | Diabetes | 0.32 |
| | Epilepsy | 0.13 |
| | Osteoporotic fracture | 0.04 |
| | Parkinson's disease | 0.08 |
| | Peripheral vascular disease | 0.14 |
| | Pulmonary hypertension or pulmonary fibrosis | 0.09 |
| | Rare pulmonary diseases | 0.12 |
| | Rare neurological conditions | 0.03 |
| | Rheumatoid arthritis or systemic lupus erythematosus | 0.14 |
| | Mental illness | 0.25 |
| | Stroke or transient ischaemic attack | 0.21 |
| | Venous thromboembolism | 0.02 |
| Region | East of England | 0.04 |
| | East Midlands | 0.01 |
| | London | 0.04 |
| | North East | 0.05 |
| | North West | 0.06 |
| | South East | 0.08 |
| | South West | 0.01 |
| | West Midlands | 0.03 |
| | Yorkshire and the Humber | 0.02 |
| Population density (people per square kilometre) | Mean | 0.05 |
| Index of Multiple Deprivation | Decile 1 (most deprived) | 0.14 |
| | Decile 2 | 0.09 |
| | Decile 3 | 0.05 |
| | Decile 4 | 0.02 |
| | Decile 5 | 0.01 |
| | Decile 6 | 0.04 |

| **Variable** | **Level** | ***d*** |
|---|---|---|
| | Decile 7<br>Decile 8<br>Decile 9<br>Decile 10 (least deprived) | 0.06<br>0.08<br>0.10<br>0.14 |
| National Statistics Socio-Economic Classification (NS-SEC) of household reference person | Higher managerial and professional occupations<br>Intermediate occupations<br>Routine and manual occupations<br>Never worked or long-term unemployed<br>Not applicable | 0.41<br>0.15<br><0.01<br>0.13<br>0.41 |
| Tenure of household | Owned outright<br>Owned with a mortgage<br>Shared ownership<br>Social rented from council<br>Other social rented<br>Private rented<br>Living rent free | 0.19<br>0.42<br>0.02<br>0.21<br>0.18<br>0.14<br>0.03 |
| Household size | 1-2 people<br>3-4 people<br>5-6 people<br>7+ people | 0.27<br>0.31<br>0.09<br>0.02 |
| Family status | Not in a family<br>In a couple family<br>In a lone parent family | 0.20<br>0.24<br>0.02 |
| Household composition | Single-adult household<br>Two-adult household<br>Multi-generational household<br>Other 3+ adults<br>Child in household | 0.30<br>0.01<br>0.02<br>0.17<br>0.34 |
| Keyworker in household | Yes<br>No | 0.16<br>0.11 |
| Household exposure to disease score | 0.0 - 20.0<br>20.1 – 40.0<br>40.1 – 60.0<br>60.1 – 80.0<br>80.1 – 100 | 0.04<br>0.03<br>0.02<br>0.06<br>0.06 |
| Household proximity to others score | 0.0 - 20.0<br>20.1 – 40.0<br>40.1 – 60.0<br>60.1 – 80.0<br>80.1 – 100 | 0.14<br>0.02<br>0.03<br>0.01<br>0.10 |

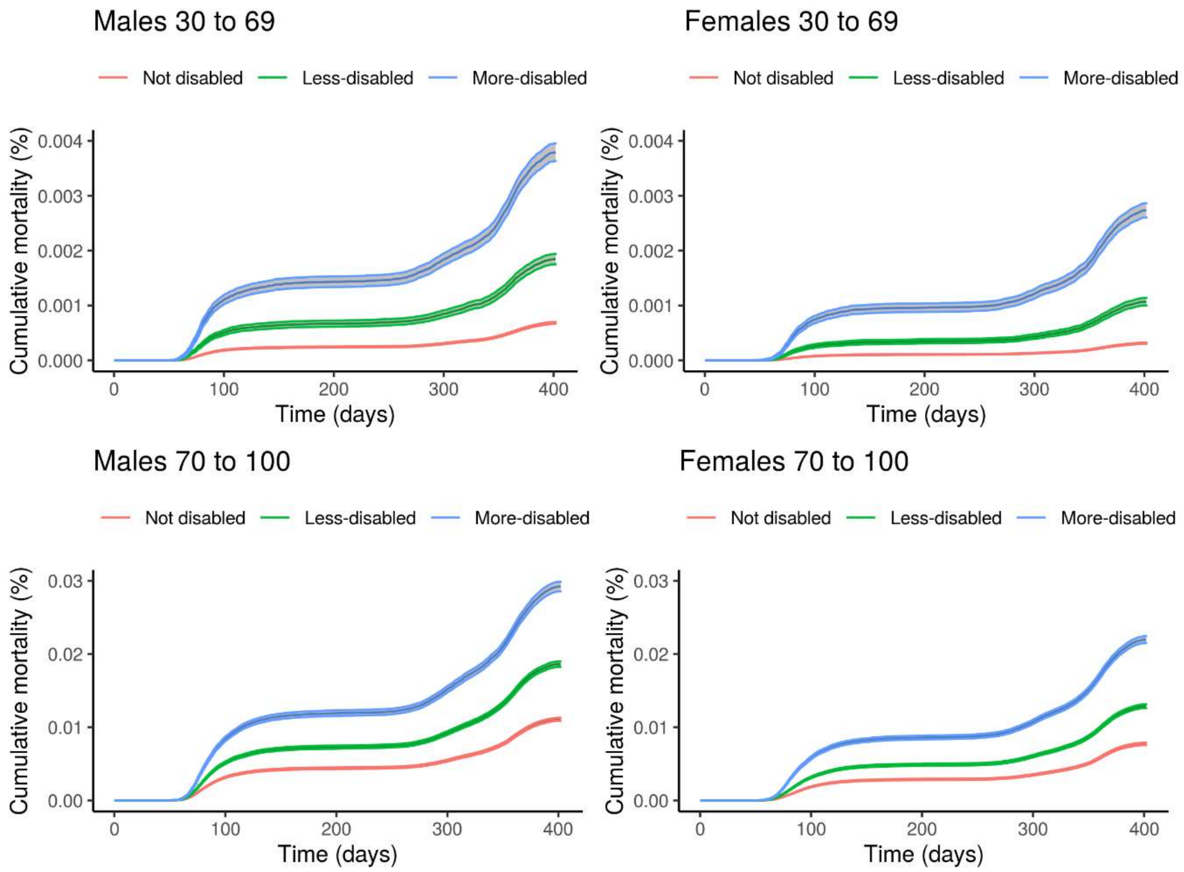


**Figure S1.** Age-adjusted cumulative COVID-19 related mortality during the outcome period (24 January 2020 to 28 February 2021) with bootstrapped 95% confidence intervals by disability status, stratified by age group (30 to 69 years and 70 to 100 years) and sex.

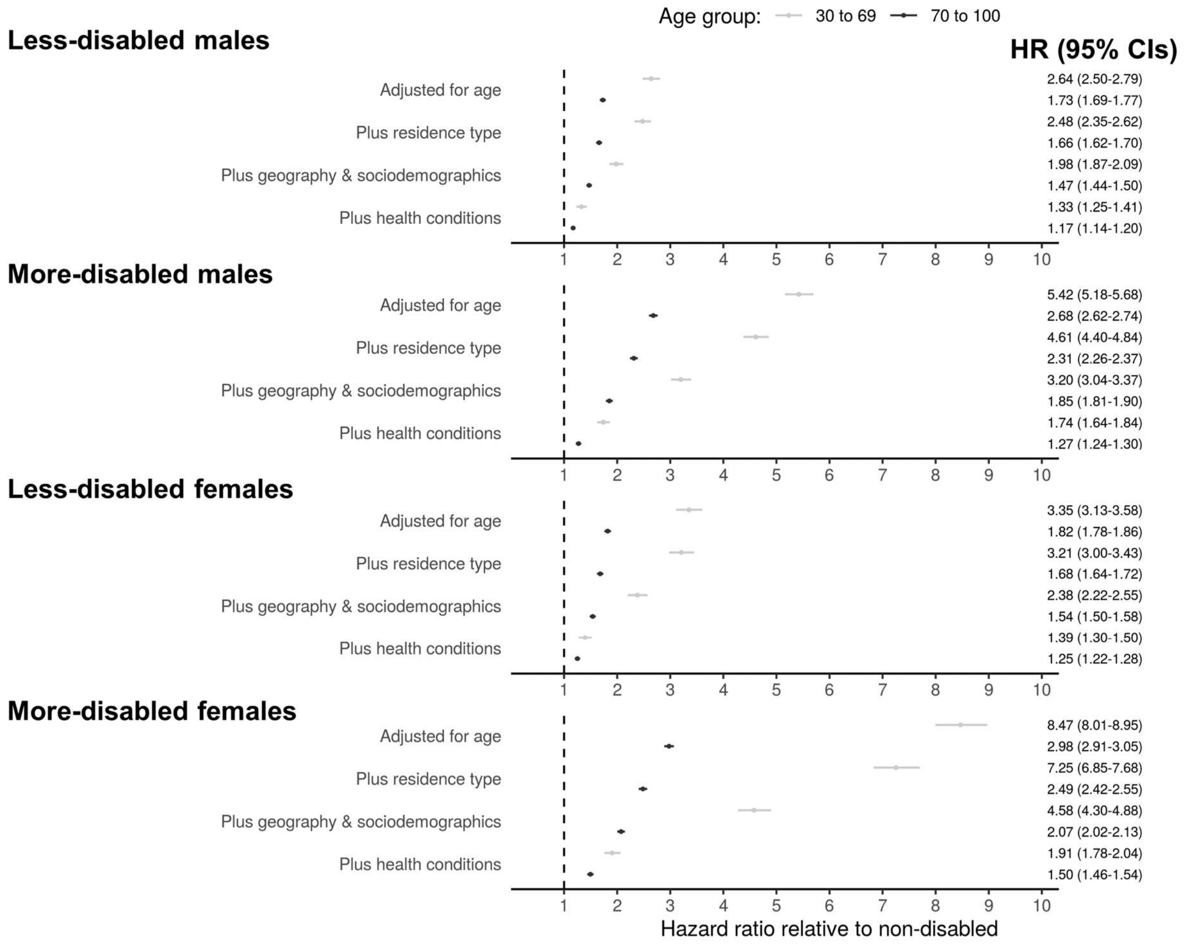


**Figure S2.** Hazard ratios for COVID-19 related mortality for less-disabled and more-disabled people compared with non-disabled people in people aged 30 to 69 years or 70 to 100 years in 2020, stratified by sex. Results obtained from Cox proportional hazards regression models adjusted for; i) age, ii) plus residence type, iii) plus local authority district, population density, area deprivation, socioeconomic status (excluding NS-SEC as this only applies to people under 75 years), ethnicity, household composition, and occupational exposure, and iv) pre-existing conditions. Error bars represent 95% confidence intervals of the hazard ratios.
